# Neurodevelopmental vulnerability in Alzheimer’s disease and frontotemporal dementia

**DOI:** 10.1101/2025.03.19.25324189

**Authors:** Perrine Laury Marie Siguier, Mélanie Planton, Bérengère Pages, Fleur Gérard, Marie Rafiq, Marie Wolfrum, Ombeline Archambault, Anise Damour, Valentine Guidolin, Pauline Pefferkorn, Lola Danet, Laurine Virchien, Eloi Magnin, Aurélie Richard-Mornas, Mathilde Sauvée, Catherine Thomas-Antérion, Servane Mouton, Mélanie Jucla, Jérémie Pariente

## Abstract

**BACKGROUND AND OBJECTIVES:** Neurodevelopmental disorders (NDDs) may influence the course of Alzheimer’s disease (AD) and frontotemporal dementia (FTD). However, prior studies have focused on specific pairs of NDDs and variants of AD/FTD, limiting generalizability. Adopting a dimensional approach to NDDs and considering the heterogeneity of AD/FTD, we investigated whether a neurodevelopmental vulnerability (DV) is associated with clinical presentation and age at onset in AD and FTD.

**METHODS:** We prospectively and consecutively recruited 84 AD/FTD participants and selected 41 matched controls. AD/FTD participants were classified into typical (amnestic AD, behavioral FTD) and focal (primary progressive aphasia, frontal and posterior variants of AD, right temporal variant of FTD, amnestic FTD) presentations. All participants underwent a neuropsychological assessment and answered a novel questionnaire on NDDs symptoms. Using k-means clustering, participants were assigned to a DV+ (with neurodevelopmental vulnerability) or a DV− (without) cluster, based on their responses on the questionnaire. This data-driven approach enabled an unbiased classification of individuals with a DV, beyond traditional diagnostic labels.

**RESULTS:** DV frequencies did not differ between the AD/FTD (18%) and control (15%) groups (χ²=.205; p=.651); and between the typical (21%) and focal (11%) subgroups (Fisher’s test, p=.184). However, in DV+ patients, symptom onset occurred 8.0 years earlier than in DV− patients (95% CI [−14, −3.0]; p = .005), with a median onset age of 58 years (IQR: 15).

**DISCUSSION:** Our findings do not support an increased risk of dementia in DV+ individuals, including in focal presentations. However, a DV would significantly hasten symptom onset. Thus, DV may act as a disease modifier and should be considered in clinical trial design, particularly for early-onset dementia. Further research is needed to elucidate the neurophysiological mechanisms linking DV to early-onset AD/FTD, with implications for precision medicine and individualized treatment strategies.

*Study registration numbers:* RnIPH 2023-71 and Research Ethics Committee file No. 2023_765

## 1. BACKGROUND

Alzheimer’s disease (AD)^a^ and frontotemporal dementia (FTD) are two neurocognitive disorders encompassing a wide variability of clinical presentations. Most frequently, AD and FTD are characterized by amnestic and behavioral symptoms, respectively. However rarer, more focal cortical neurodegeneration processes can also lead to initial linguistic, visuospatial or emotional manifestations ^1–4^. Likewise, though AD and FTD usually affect adults in their later years, they can also arise before the age of 65 ^5^. This great heterogeneity of symptoms and age at onset is only partially explained by physiopathology, and might result from the interaction between molecular and individual factors ^6^.

These individual factors encompass neurodevelopmental disorders (NDDs), a group of syndromes emerging during childhood and impairing personal, social, academic, and occupational spheres ^7^. However, functional particularities associated with NDDs frequently persist into adulthood ^8–10^. Thus, NDDs modify life-span neurocognitive trajectories, to the extent that their interaction with neurodegenerative disorders has even been suggested ^11,12^.

More specifically, NDDs would confer a higher risk of developing dementia (e.g., ^13,14^). Besides, some studies showed that NDDs are more frequent in focal cortical variants (Fv) of AD and FTD ^15,16^. Finally, they have also previously been associated with a younger age at onset of dementia (e.g., ^17,18^). All together, these results suggest that NDDs could influence the course of neurodegenerative diseases (for a review, see ^19^). Nevertheless, the majority of these investigations exclusively focused on specific variants of AD and FTD, thus eluding a large part of their presentations. They also targeted distinct NDDs, whereas recent studies show the importance of adopting a dimensional approach, rather than a categorical one, to account for the overlap of these syndromes (e.g., ^20,21^).

One way of achieving this is by focusing on symptoms rather than on labels of NDDs, which is one of the advantages provided by questionnaires ^22,23^. Contrary to other methods such as screening medical records, these tools also enable a rapid, standardized collection of information, that can be confirmed by a third party. However, few retrospective questionnaires exist and most of them screen a specific category of NDD. Moreover, in the absence of validation, a history of NDD is sometimes determined on the basis of an arbitrary threshold ^19,24^.

In this monocentric prospective study, we used a novel retrospective heteroquestionnaire covering a wide range of symptoms characterizing NDDs to screen for a neurodevelopmental vulnerability in populations with AD or FTD and a group of controls. The presence of a neurodevelopmental vulnerability was determined by means of a data-driven approach, the interest of which has already been illustrated in the fields of NDDs and neurodegenerative diseases.

We aimed at comparing the frequencies of a neurodevelopmental vulnerability in the different variants of AD and FTD and in cognitive ageing; as well as to evaluate the association between a neurodevelopmental vulnerability and the age at onset of AD and FTD. The following hypotheses were formulated. First, a neurodevelopmental vulnerability would be more frequent in AD or FTD than in cognitive ageing. Second, the frequency of a neurodevelopmental vulnerability in Fv of AD and FTD would be higher than the frequency in typical variants. Third, a neurodevelopmental vulnerability would be associated with earlier onset of AD and FTD.

## 2. METHODS

This study followed the Strengthening the Reporting of Observational Studies in Epidemiology (STROBE) reporting guideline.

### 2.1. Standard Protocol Approvals, Registrations, and Participant Consents

Participants were prospectively recruited from October 2023 to July 2024. All provided written informed consent and the study was approved by local institutional review boards (RnIPH 2023-71 and Research Ethics Committee file No. 2023_765). The protocol was in accordance with the Helsinki Declaration. Participants had French as first language and no comorbid illness altering their cognition.

#### 2.1.1. Participants with AD/FTD

Among neurocognitive disorders, this study focuses on AD and FTD, characterized by primary impairments in cognition and/or behavior that are not attributable to an acquired brain event. The choice to focus on AD and FTD was to limit the heterogeneity of outcome measures and to facilitate comparability with previous studies ^19^. Participants with AD or FTD (hereafter called AD/FTD group) were consecutively recruited in the memory clinic of the Neurology Department of Purpan University Hospital (Centre Mémoire de Ressources et de Recherche, Toulouse, France). AD was diagnosed following the criteria of Dubois et al. (2014) ^1^. Variants of FTD were diagnosed following criteria of Gorno-Tempini et al. (2011), Rascovsky et al. (2011) and Ulugut et al. (2024) ^2–4^. Participants with AD/FTD were required to have a MMSE score ≥18 ^25^ ensuring their ability to complete the neuropsychological assessment. They were sub-classified as having typical or focal cortical variants of AD/FTD depending on the initial symptoms they experienced. This implied a double-blinded procedure: four neurologists (FG, MR, MW, JP) classified patients on the basis of clinical features; while a neuropsychologist (BP) classified them based on scores of the first previous neuropsychological assessment available. Typical variants (Tv) were amnestic AD (aAD) and behavioral FTD (bvFTD). Focal cortical variants (Fv) of AD were logopenic primary progressive aphasia (lvPPA), but also frontal, and posterior cortical atrophy variants. Fv of FTD were semantic PPA (svPPA), non-fluent/agrammatic PPA (nfvPPA) and right temporal variant (rtvFTD). Initial amnestic presentation of FTD was categorized as an unclassified Fv (cf. Supplementary Table 1 for detailed samples sizes).

#### 2.1.2. Control participants

The control group encompasses participants without cognitive complaints, and is limited to participants with a MMSE score indicating no general cognitive impairment relative to their education level ^25^. Forty-one individuals were randomly selected out of a data base of control participants who followed the present protocol, so that the mean age, the number of years of schooling and the sex ratio were comparable with those from the AD/FTD group ^b^.

### 2.2. Neurodevelopmental vulnerability questionnaire

All participants answered a novel questionnaire evaluating life-span NDDs symptoms (cf. Supplementary Figure 1 for English version; Original French version upon reasonable request). The questionnaire screens for NDDs traits, rather than providing a clinical diagnosis. It was developed by SM in collaboration with the GRECO-GREDEV (working group for the assessment of NDDs in adults), a French multidisciplinary team of experts including neurologists, psychiatrists, neuroscientists, neuropsychologists and speech and language therapists.

The questionnaire includes 4 items of anamnesis (scored 0 if no element is reported, 1 if at least one is reported); followed by 46 items with a Likert scale from 0 to 5 (27 for childhood, e.g., “Did you find it hard to sit still? Were you told you were unruly and restless?”; 19 for adulthood, e.g., “Do you have difficulties with spelling?”). Higher scores reflect more frequent or severe difficulties. The experimenter read all items out loud and collected participants’ answers. In the AD/FTD group, but not in the control group, the questionnaire was filled according to a consensus between the patient and their relatives.

### 2.3. Neuropsychological assessment

Participants underwent a cognitive assessment evaluating language [BECS-GRECO confrontation naming ^26^; ECLA16^+^ irregular words dictation ^27^], executive functioning [“S” lexical fluency, Go–No Go and Conflicting instructions from the FAB ^28^], visuospatial processing [Rey-Osterrieth figure copy ^29,30^] and anterograde memory [Rey-Osterrieth figure delayed recall ^30^].

### 2.4. Statistical analysis

Analyses were performed using R Studio and Jamovi. Firstly, an exploratory factor analysis (EFA) was performed on the 27 items of the childhood part of the questionnaire. Adulthood scores were not included in the EFA to prevent potential early neurodegenerative symptoms to be mistaken for NDDs traits. Secondly, based on the standardized factors extracted from the EFA, a k-means clustering algorithm with 100 random starting values was used to partition all participants in k=2 clusters ^c^. The number of clusters was defined *a priori* based on the assumption that the two clusters would reflect the presence or the absence of a neurodevelopmental vulnerability, respectively. Clusters were characterized through comparison of mean factors scores and through comparison of rescaled scores on the anamnesis, the childhood and the adulthood parts of the questionnaire. Rescaled scores were calculated as the percentage of the maximum possible score in each part of the questionnaire. Thirdly, the frequencies of control participants and of patients with typical or focal cortical variants were compared in each cluster using Chi-square and Fisher’s exact tests. Finally, age at onset and at diagnosis of AD/FTD were compared in patients from the two clusters using Student and Mann-Whitney tests.

### 2.5. Data Availability

Data are available from the corresponding author on reasonable request.

## 3. RESULTS

### 3.1. Participants

A total of 125 participants (51 with AD, 33 with FTD, 41 controls) were included in the analyses (cf. Figure 1). Among subjects with AD/FTD, 56 (67%; 38 AD, 18 FTD) were classified in the Typical variants (Tv) subgroup and 28 (33%; 13 AD, 15 FTD) were classified in the focal cortical variants (Fv) subgroup. The three subgroups were comparable in terms of age, level of education and sex ratio but patients had lower MMSE scores. Forty-six percent of participants with AD/FTD had a familial history of neurodegenerative diseases (n=39). Eight individuals had genetic AD (n=2) and FTD (n=6). Patients were accompanied by their spouses (71% of cases, n=60), their children (14%, n=12) or other relatives (15%). The rescaled scores of the anamnesis, the childhood and the adulthood parts of the questionnaire were similar among the subgroups. All the neuropsychological assessment indicators, except the central coherence index of the Rey figure, showed impaired performances for participants with AD/FTD compared to the control group (cf. Table 1). Rey figure copy score and recall time could not differentiate groups once p-values were adjusted.

**Figure 1:**
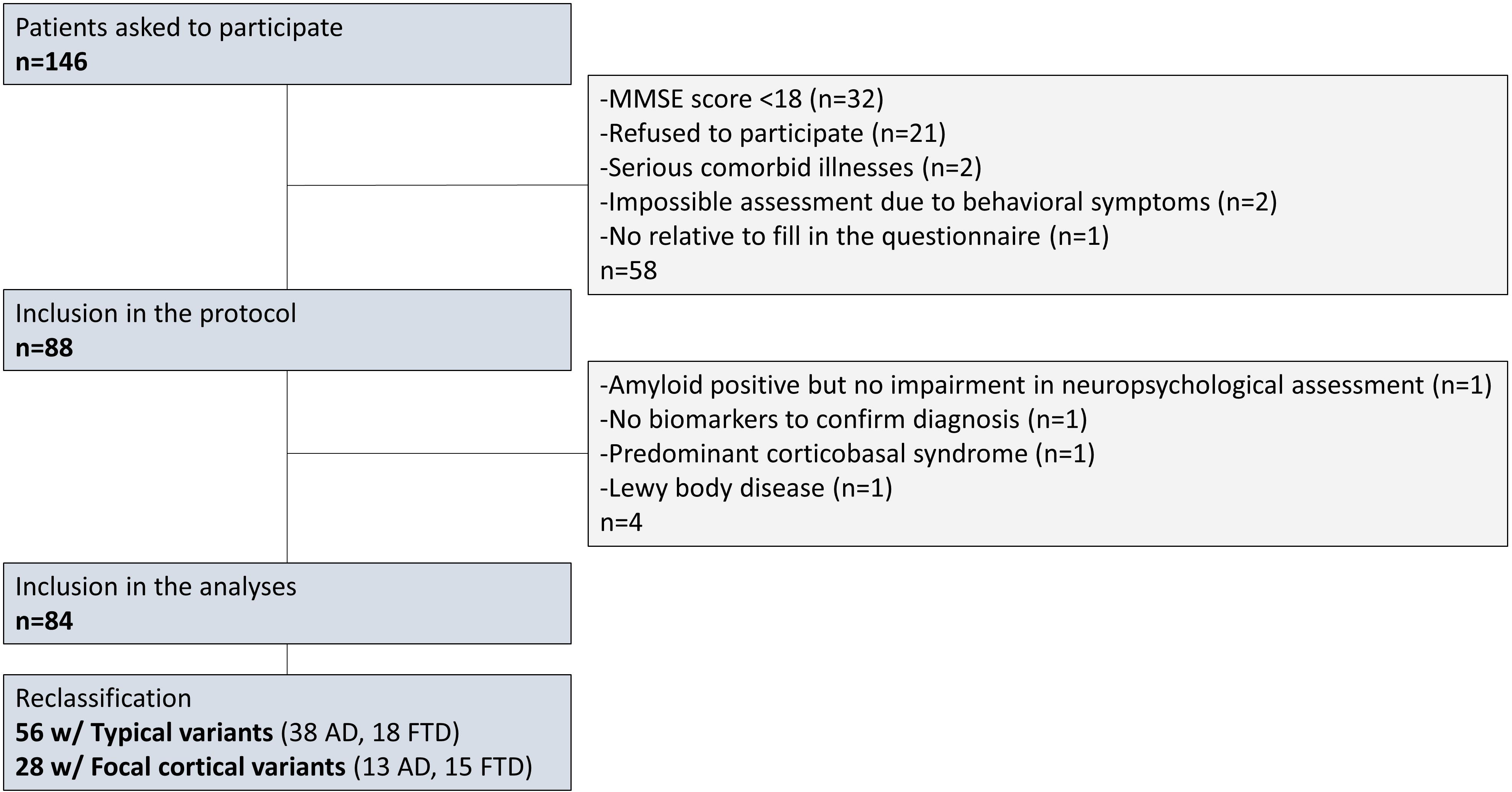
Flowchart of AD/FTD participants. The 21 patients who refused to participate did not statistically differ from included patients on any of the following variables: age, level of education, MMSE, sex ratio, proportions of AD/FTD, proportions of Tv/Fv, proportions of genetic cases, age at onset, age at diagnosis, time between first symptoms and diagnosis, time since diagnosis (cf. Supplementary Table 2).

**Table 1:** Demographical and clinical descriptions of subgroups. Item A) “Have you been diagnosed with a neurodevelopmental disorder? If so, which one(s)?”. Item B) “Have you repeated a year and/or been in an adapted class?”. Item C) “Did you need help learning to read, write, count or draw in primary school?”. Item D) Did you receive medical or paramedical support, such as: speech and language therapy, occupational therapy, psycho-motor therapy, psychologist, child psychiatrist, neuropsychologist?”. *One-way ANOVA for independent samples. ^†^Kruskal-Wallis tests, df=2. ^‡^ Kruskal-Wallis tests with Bonferroni correction. ^§^Post-hoc Dunn’s test with Holm correction showed p-adjusted < .005 for control – focal. ^¶^Post-hoc Dunn’s test with Holm correction showed p-adjusted < .005 for control – typical. ^#^Post-hoc Dunn’s test with Holm correction showed p-adjusted < .005 for focal – typical.

|  |  | Subgroups |  |  | Statistic | p-value |
| --- | --- | --- | --- | --- | --- | --- |
|  |  | Typical<br>(n=56) | Focal<br>cortical<br>(n=28) | Control<br>(n=41) |  |  |
| Age (years) | Mean (sd) | 72.0 (8.4) | 70.6 (8.9) | 70.2<br>(7.9) | F=.611,<br>df1=2,<br>df2=122 | .544* |
|  | Min/Max | 45.0/88.3 | 52.3/85.4 | 53.0/94.0 |  | - |
| Level of<br>education (years<br>of schooling) | Median<br>(IQR) | 11.0 | 12.0 | 14.0 | $\chi^2=4.18$ | .124<sup>†</sup> |
|  | Min-Max | 5-20 | 5-20 | 5-18 |  | - |
| MMSE score | Median<br>(IQR) | 23.0 (4.0) | 24.5 (4.0) | 29.0<br>(1.0) | $\chi^2=109.75$ | <.05<sup>†§¶</sup> |
|  | Min-Max | 18-28 | 18-29 | 26-30 |  | - |
| Sex ratio | Number of<br>males (% in<br>subgroup) | 30 (54) | 15 (54) | 21 (51) | - | .970 |
| Questionnaire<br>Anamnesis score<br>(/4) | Median<br>(IQR) | 0 (1) | 0 (1) | 0 (1) | $\chi^2=1.911$ | .385<sup>†</sup> |
|  | Min-Max | 0-2 | 0-3 | 0-3 |  |  |
| Item A: NDD<br>diagnosis | No. of score<br>1 (%) | 1 (2) | 3 (11) | 0 (0) | - | - |
| Item B: adapted<br>education | No. of score<br>1 (%) | 18 (32) | 7 (28) | 19 (46) | - | - |
| Item C: help for<br>learning | No. of score<br>1 (%) | 6 (11) | 1 (4) | 3 (7) | - | - |
| Item D: medical<br>support | No. of score<br>1 (%) | 1 (2) | 2 (7) | 1 (2) | - | - |
| Questionnaire<br>Childhood<br>rescaled score<br>(%) | Median<br>(IQR) | 6.3 (16) | 5.9 (10) | 7.4 (10) | $\chi^2=3.061$ | .216<sup>†</sup> |
|  | Min-Max | 0-38 | 0-20 | .7-27 |  |  |
| Questionnaire<br>Adulthood<br>rescaled score<br>(%) | Median<br>(IQR) | 10 (17) | 10 (13) | 13 (11) | $\chi^2=.578$ | .749<sup>†</sup> |
|  | Min-Max | 0-42 | 0-32 | 0-62 |  |  |
| Confrontation<br>naming score<br>(/40) | Median<br>(IQR) | 36.0 (4.3) | 34.0 (8.0) | 38.0<br>(2.0) | $\chi^2=32.21$ | <.05<br>‡§¶# |
| Confrontation<br>naming time (s) | Median<br>(IQR) | 102.0<br>(63.3) | 154.5<br>(132.3) | 73.5<br>(27.0) | $\chi^2=67.09$ | <.05<br>‡§¶ |
|  | Missing | 0 | 2 | 0 |  |  |
| Dictation score<br>(/10) | Median<br>(IQR) | 5 (5) | 5 (3) | 8 (3) | $\chi^2=27.65$ | <.05<br>‡§¶ |
|  | Missing | 0 | 1 | 0 |  |  |
| Dictation time (s) | Median<br>(IQR)<br>Missing | 71.0<br>(36.5)<br>1 | 83.5<br>(47.3)<br>4 | 53.0<br>(14.0)<br>0 | $\chi^2=42.35$ | **<.05**<br>‡§¶ |
| Fluency (letter S) | Median<br>(IQR)<br>Missing | 8.0 (4.5)<br>1 | 6.0 (7.3)<br>0 | 12.0<br>(6.3)<br>0 | $\chi^2=38.14$ | **<.05**<br>‡§¶ |
| Go–No Go score (/3) | Median<br>(IQR)<br>Missing | 2.0 (2.0)<br>1 | 2.5 (2.0)<br>0 | 3.0 (0)<br>0 | $\chi^2=31.26$ | **<.05**<br>‡§¶ |
| Conflicting instructions score (/3) | Median<br>(IQR) | 2.0 (2.0) | 2.5 (2.0) | 3.0 (0) | $\chi^2=30.65$ | **<.05**<br>‡§¶ |
| Rey copy score (/36) | Median<br>(IQR)<br>Missing | 26.0<br>(12.0)<br>1 | 27.3 (9.6)<br>0 | 28.0<br>(5.1)<br>0 | $\chi^2=8.92$ | .416<sup>‡</sup> |
| Rey copy time (s) | Median<br>(IQR)<br>Missing | 262 (164)<br>3 | 278 (139)<br>2 | 147 (62)<br>0 | $\chi^2=55.27$ | **<.05**<br>‡§¶ |
| Rey delayed recall score (/36) | Median<br>(IQR)<br>Missing | 3.8 (8.5)<br>2 | 5.5 (11.3)<br>0 | 14.0<br>(6.1)<br>0 | $\chi^2=57.63$ | **<.05**<br>‡§¶ |
| Rey delayed recall time (s) | Median<br>(IQR)<br>Missing | 81 (54)<br>10 | 103 (73)<br>6 | 101 (50)<br>0 | $\chi^2=6.54$ | 1.00<sup>‡</sup> |
| Rey Central Coherence Index | Median<br>(IQR)<br>Missing | 1.33 (0.8)<br>2 | 1.26 (0.8)<br>0 | 1.33<br>(0.5)<br>0 | $\chi^2=.59$ | 1.00<sup>‡</sup> |

### 3.2. Data-driven analysis

#### 3.2.1. Exploratory factorial analysis

Bartlett’s test of sphericity (χ²=957, ddl=253, p<.001) indicated that the correlation structure of the EFA was adequate for factor analyses. Items 5, 18, 21 and 25 were removed from EFA because of inappropriate sampling adequacy (Kaiser-Meyer-Olkin test values <.5; general KMO = .692, cf. Suppl. Table 3). Principal axis extraction was used for its applicability on non-normal distributions. Oblimin rotation was chosen to allow collinearity between factors. The solution offering the best compromise between interpretability of factors and accounted variance (44.5%) was yielded by a parallel analysis with a cut-off point of .4 and six factors. The results of this analysis are presented in Suppl. Table 4. The factors were interpreted according to the cognitive domains addressed in the contributing questions. Correlations between factors are presented in Suppl. Table 5.

#### 3.2.2. K-means clustering

Cluster 1 and cluster 2 encompassed 21 (16.8%) and 104 (83.2%) individuals, respectively (cf. Suppl. Figure 2). Compared to cluster 2, cluster 1 was characterized by significantly higher mean values in 4 out of 6 factors (cf. Suppl. Table 6). It also had significantly higher rescaled scores in each part of the questionnaire (cf. Table 2). These results suggested that Cluster 1 included participants with a neurodevelopmental vulnerability (DV). Therefore, cluster 1 and cluster 2 will hereafter be named “DV+” and “DV-”, respectively. Clusters did not differ in terms of level of education, MMSE score and sex ratio, but mean age was lower in the DV+ than in the DV-cluster.

**Table 2:** Demographical description of clusters. Fv=Focal cortical variants of AD/FTD, Tv=Typical variants of AD/FTD. *Mann-Whitney test. ^†^Independent samples T-test. ^‡^Fisher’s exact test. ^§^Unilateral Chi-square test, alternative hypothesis: P(controls)<P(patients).

|  |  | Clusters |  | Statistic | p-value |
| --- | --- | --- | --- | --- | --- |
|  |  | DV+<br>(n=21) | DV-<br>(n=104) |  |  |
| Questionnaire Anamnesis score (/4) | Median (IQR)<br>Min-Max | 1 (1)<br>0-3 | 0 (1)<br>0-2 | U=488 | **<.001\*** |
| Item A: NDD diagnosis | No. of score 1 (%) | 3 (14) | 1 (1) | - | - |
| Item B: adapted education | No. of score 1 (%) | 14 (67) | 30 (29) | - | - |
| Item C: help for learning | No. of score 1 (%) | 6 (29) | 4 (4) | - | - |
| Item D: medical support | No. of score 1 (%) | 4 (19) | 0 (0) | - | - |
| Questionnaire Childhood rescaled score (%) | Median (IQR)<br>Min-Max | 24 (7.4)<br>17-38 | 5.6 (8.2)<br>0-22 | 13.0 | **<.001\*** |
| Questionnaire Adulthood rescaled score (%) | Median (IQR)<br>Min-Max | 22 (8.4)<br>3.2-62 | 9.0 (12)<br>0-36 | 368.0 | **<.001\*** |
| Age (years) | Mean (sd)<br>Min-Max | 66.6 (10.1)<br>45.0-87.4 | 72.1 (7.71)<br>52.1-94.0 | t=-2.80 | **.006<sup>†</sup>** |
| Level of education (years of schooling) | Median (IQR)<br>Min-Max | 11 (5)<br>5-20 | 12 (4)<br>5-20 | U=897 | .191*<br>- |
| MMSE score | Median (IQR)<br>Min-Max | 25.0 (3.00)<br>18-30 | 25.5 (6.00)<br>18-30 | U=1076 | .918*<br>- |
| Sex ratio | Number of males (% in cluster) | 14 (66.7) | 52 (50) | - | .231<sup>‡</sup> |
| Patients/Controls ratio | Patients/Controls (% patients in cluster) | 15/6<br>(71) | 69/35<br>(66) | $\chi^2 = .205$ | .651<sup>§</sup> |

### 3.3. Is a DV associated with AD/FTD?

Of the 84 AD/FTD participants and the 41 controls, 17.9% and 14.6% were classified in the DV+ cluster, respectively. In other words, the proportions of AD/FTD and of control participants in the two clusters were comparable (χ²= .205; p=.651) (cf. Table 2). Thus, no association was found between groups and clusters.

### 3.4. Is a DV associated with focal cortical variants of AD/FTD?

The DV+ cluster regrouped 10.7% of the 28 participants with Fv of AD/FTD and 21.4% of the 56 participants with Tv of AD/FTD. These proportions were not statistically different in the two clusters (p=.184) (cf. Table 3). Thus, no association was found between subgroups and clusters.

**Table 3:**
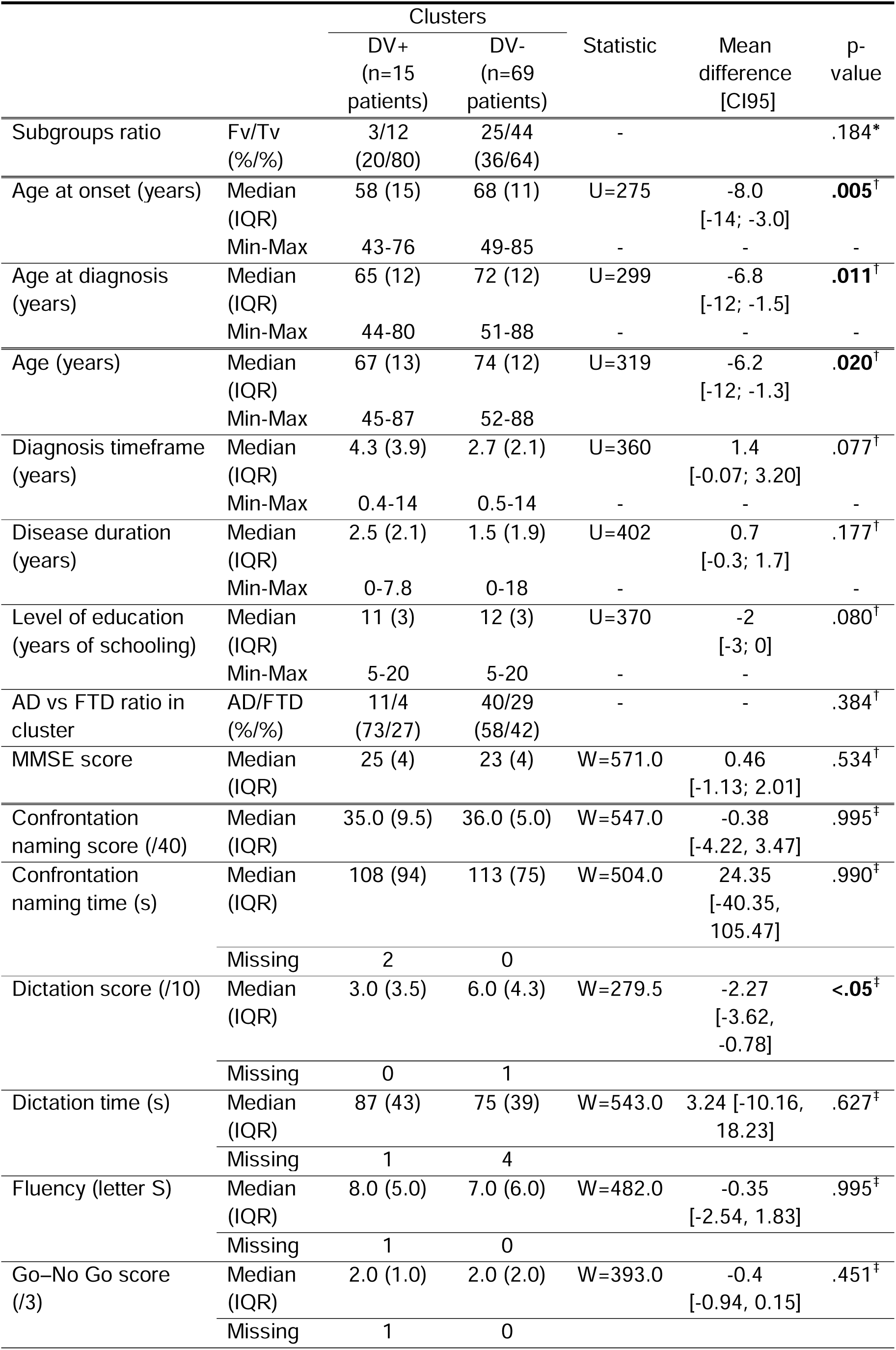
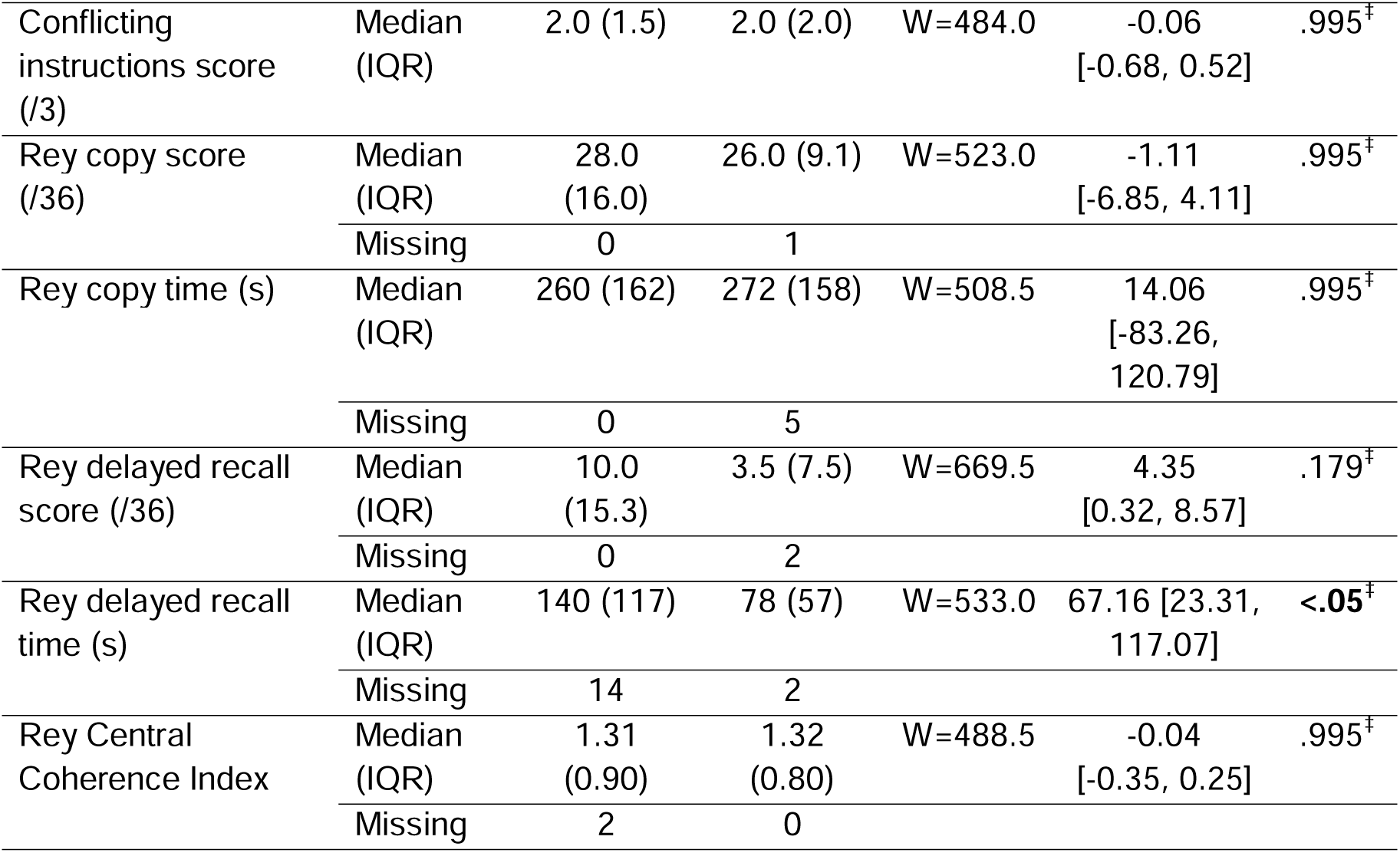
Characterization of AD/FTD participants in clusters. Diagnosis timeframe corresponds to the time between symptoms onset and diagnosis. Disease duration corresponds to the time between diagnosis and inclusion in the study. *Note*: One missing value for age at onset and diagnosis delay. *Fisher’s exact test. ^†^Bilateral Mann-Whitney test. ^‡^Bilateral Mann-Whitney test with FDR correction.

### 3.5. Is a DV associated with earlier onset AD/FTD?

Patients in the DV+ cluster had a significant earlier age at onset (i.e., age at first symptoms; mean difference = −8.0 years, 95CI [-14;-3.0], p=.005) and at diagnosis (mean difference = −6.8, 95CI [-12;-1.5], p=.011) (Figure 2, detailed statistical parameters in Table 3), in spite of comparable diagnosis timeframe (estimated as time between first symptoms and the diagnosis) and duration of AD/FTD (estimated as time between the diagnosis and the inclusion in the study). Notably, proportions of cases with AD and FTD were equivalent in the two clusters. Likewise, genetic forms of AD (n=2) and FTD (n= 6) were not more frequent than expected in the DV+ cluster (right-tailed Binomial test, x=2, n=8, expected proportion=.18, observed proportion=.25, 95CI [0.046; 1.000], *p-value*=.432). No association was found between clusters and a familial history of neurodegenerative diseases (χ² = 5.06×10^-31^, df = 1, *p-value* = 1). Besides, the age difference previously found between clusters (Table 2) was mediated by patients. Indeed, this difference persists when only patients are considered (Table 3), and is not found in controls (U=88, mean difference=-2.0, 95CI [-11;5.0], p=.542).

**Figure 2:**
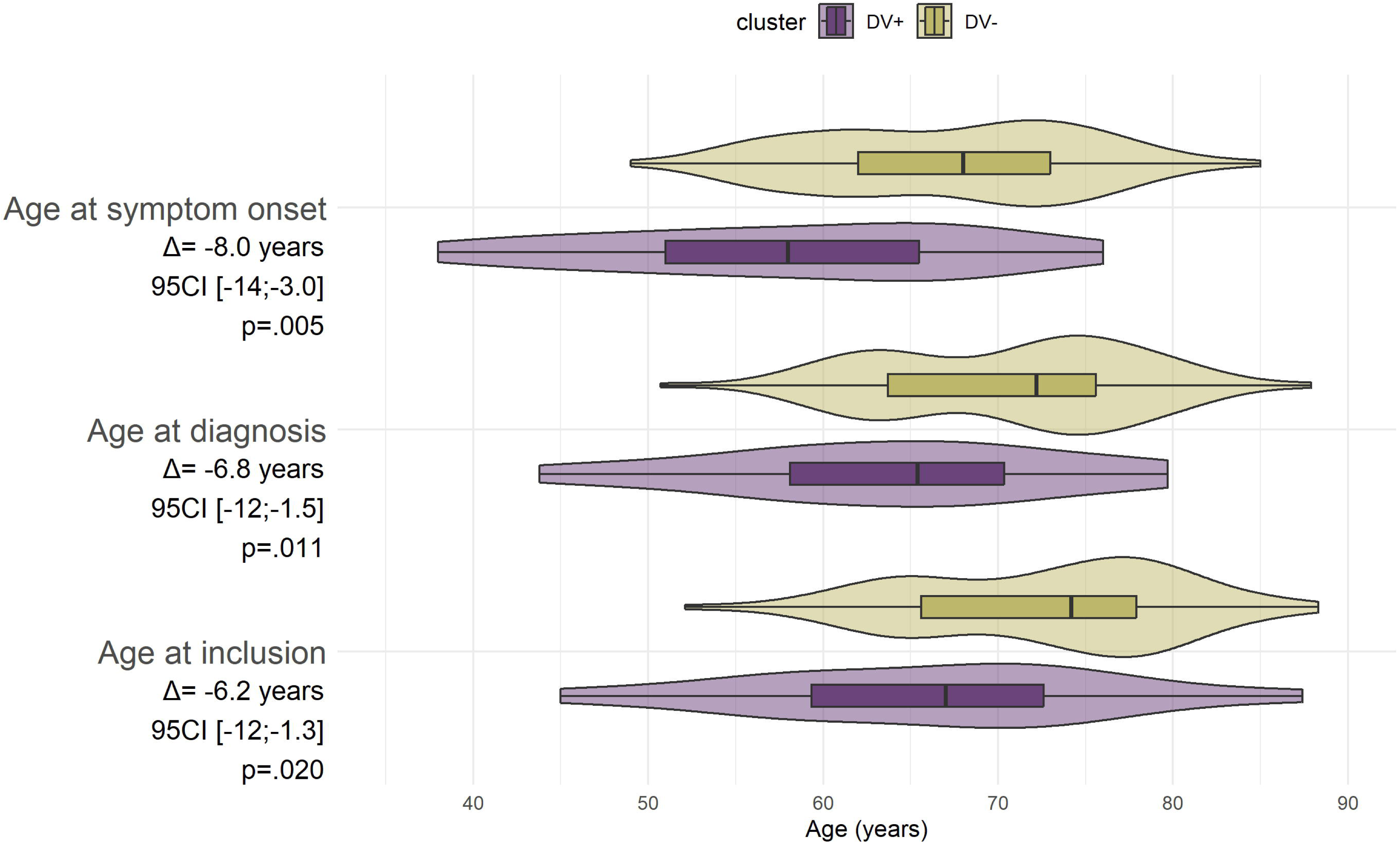
Participants with AD/FTD and a developmental vulnerability showed earlier onset of disease. The letter “Δ” refers to the mean difference between the median in the DV+ cluster and the median in the DV-cluster. The “95CI” is the 95% confidence interval of the mean difference.

## 4. DISCUSSION

We identified individuals with a developmental vulnerability (DV) using a data-driven analysis conducted on a novel questionnaire. We showed that participants with AD/FTD and a DV experience symptoms of AD/FTD in average 8 years earlier than those without a DV. More precisely, in our sample, the onset of AD/FTD with a DV occurred before the age of 60 in the majority of participants. In this regard, a DV seems associated with early-onset AD/FTD ^5^.

AD/FTD was also diagnosed approximately 7 years earlier in participants with a DV, which explains the significant difference in age at inclusion found between the two clusters. Indeed, the time between onset and diagnosis and between diagnosis and inclusion was comparable between the two clusters. Younger age at onset and at diagnosis was not mediated by participants with FTD, with genetic forms of AD/FTD or with familial history of neurodegenerative diseases, as their repartition into the two clusters was homogeneous. Likewise, the clustering of participants with AD/FTD was not driven by their cognitive functioning level, as showed by their similar MMSE and neuropsychological scores in the two clusters. As a matter of fact, participants with AD/FTD of the two clusters only differed on their dictation scores and on the complex figure delayed recall time. DV+ patients had a lower dictation performance and spent more time on the figure recall. In the absence of an exhaustive neuropsychological assessment, it seems arduous to determine if these differences arise from neurodegenerative processes or from complex neurodevelopmental interactions. Yet, it appears unlikely that these disparities biased results towards the observed difference in age at onset between clusters.

More surprisingly, we did not find any difference in the level of education between clusters, considering, or not, control participants. One could have expected individuals of the DV+ cluster to display lower academic achievements than their counterparts, as has previously been mentioned (e.g., ^7,8^). This could be explained by the fact that we compared the number of years of education corresponding to the highest qualification obtained, regardless of if it was a vocational or a general education. Nevertheless, this relationship between NDDs and academic achievements may not be a general rule as it seems to be modulated by other factors (e.g., executive functioning) and to vary with age ^31^. It could also reflect a selection bias, with studies reporting this relationship focusing on individuals seeking formal diagnosis because of more severe impairments and thus, academic struggles. The fact that individuals in the DV+ cluster achieved the same level of education as their counterparts also underlines that educational level should not be used as a sole proxy for NDDs. This further emphasizes the importance of multidimensional approaches, including questionnaires, in the detection of NDDs.

Likewise, males were not predominant in the DV+ cluster, despite being frequently described as more affected by NDDs ^7,32^. In fact, the questionnaire reflects symptoms of NDDs, rather than diagnoses. In this way, observing that males and females report the same amount of symptoms of NDDs corroborates the assumption that the biased sex ratio in NDDs would mostly reflect differences in diagnosis criteria and NDDs screening ^33^. This focus on NDDs traits and not on diagnosis is also likely to explain the relatively high proportions of individuals with a childhood DV in our sample (17.9% of AD/FTD participants and 14.6% of controls) compared to, for example, the NDDs prevalence of 8.56% reported by Zablotsky and colleagues in a US sample of children aged 3–17 years ^34^. However our finding seems plausible as worldwide, the estimated prevalence of NDDs could fluctuate between 4.70 and 88.50%, depending on methodological and sociocontextual aspects ^35^.

Notably, the proportion of individuals classified as DV+ did not differ between patients and controls, or between patients with Fv and Tv of AD/FTD. In this regard, our study does not support the idea that a DV would be associated with a greater risk of dementia ^13,14,18^. While this may be due to our reduced sample size, it is important to note that previous studies reporting this association focused on specific pairs of NDDs and neurocognitive disorders. This too could explain why, considering the global spectrum of NDDs, we did not replicate these results: this association could exist only between certain NDDs and neurocognitive disorders. This is particularly applicable when investigating the link between NDDs and focal variants of AD/FTD, where NDDs would influence AD/FTD’s expression towards alterations of shared phenotype ^15,16,36,37^. More precisely, an increased prevalence of learning disorders─especially in written language─ has been reported in patients with PPA and their first-degree relatives, compared to typical variants of AD and FTD ^16,36,37^. Besides, mathematical and/or visuospatial learning disorders were shown to be more frequent in PCA than in amnestic AD, lvPPA and the general population. Nonetheless, one cannot exclude that neither typical nor focal cortical neurocognitive disorders are favored by a history of NDD, as several works actually failed to show any association (e.g., ^24,38–41^). For example, the association between learning disorders and PPA was not found in a sample of Brazilian Portuguese native speakers, suggesting that it could be language specific ^38^. Using two-sample Mendelian Randomization, another study detailed only limited evidence of a causal effect of genetic liability to attention deficit hyperactivity disorder (ADHD) or autism spectrum disorder (ASD) on AD; and did not find a causal effect of AD on risk of ADHD or ASD ^40^.

We acknowledge that this study has several limitations. First, our sample of participants with AD/FTD was recruited in a third-referral consultation. Replication in a wider variety of centers would ensure the generalizability of the findings. Second, the questionnaire we employed is retrospective and has yet to be validated. Nonetheless, the fact that it was developed by experts of NDDs lifespan trajectories should at least partly insure its face validity ^42^. We also tackled potential long-term memory biases in participants with AD/FTD by asking a close relative to confirm and modulate answers on the questionnaire. Moreover, the distinction between individuals with and without DV was made through a data-driven methodology, thus eliminating the potential bias associated with an a priori defined threshold score. Third, this data-driven methodology itself has some drawbacks, as the interpretability of the factors extracted from an EFA is not straightforward ^43^. However here, EFA was only used to reduce data dimensionality, and identifying the relationships between items was outside the scope of our objectives. The future validation of the questionnaire should nevertheless imply a confirmatory factorial analysis. Finally, we cannot exclude that the association between a DV and an early-onset AD/FTD does not reflect the influence of a DV on the course of AD/FTD; but rather that other factors predisposing for an earlier-onset neurocognitive disorder also cause impairments during childhood. Indeed, neurodevelopmental effects of AD and FTD have been suggested ^11,44,45^, and for these reasons bilateral interactions should not be ruled out either. Finger and colleagues ^44^ have for example suggested that antagonist pleiotropy could be at stake in young MAPT carriers, where a mutation would be beneficial in early life, but detrimental later. Here on the contrary, individuals having an early neurodevelopmental vulnerability appeared more disadvantaged when facing neurodegenerative processes, which is coherent with the Matthew principle ^46^. This pattern is likely to arise from joint genetic and environmental influences undermining the brain’s resilience, that is to say « [its] capacity to maintain cognition and function with aging and disease » ^47^. Deciphering more precisely if a DV is associated with a different brain reserve, a different cognitive reserve or a different brain maintenance will require further longitudinal neuroimaging studies.

To conclude, no association between a DV and typical or focal variants of AD/FTD was found in this study. Nevertheless, it builds on previous results showing that a DV is associated with early-onset AD/FTD, while extending this conclusion beyond specific NDDs. This dimensional approach of NDDs was enabled by the use of a data-driven analysis of a novel questionnaire focusing on symptoms of NDDs across their whole spectrum, rather than on categorical diagnoses ^21,22^. The heterogeneity of presentations of AD and FTD was also accounted for, thanks to a prospective and consecutive recruitment avoiding selection biases.

These findings demonstrate the importance of detecting neurodevelopmental impairments in order to provide a personalized precision medicine. This includes early-detection, differential diagnosis but also prevention of neurodegenerative diseases, and could extend to predicting therapies outcomes. As such, DVs might be viewed as disease modifiers that should be considered in the design of clinical trials. Perspectives include unraveling the neural bases underlying the link between DVs and earlier-onset neurocognitive disorders, especially longitudinally. This seems crucial in order to predict neurodegenerative trajectories of individuals with a DV^46,48,49^.

## Supporting information

STROBE_checklist_case-control

## Data Availability

All data produced in the present study are available upon reasonable request to the authors

## Acknowledgements

We wish to thank all the members of the GREDEVad (Groupe de réflexion sur l’évaluation des troubles neurodéveloppementaux de l’adulte) of the GRECO (Groupe de réflexion sur l’évaluation cognitive) for their contribution to this study. We acknowledge Ms. Caroline Court and Ms. Nathalie Gravier for their invaluable help in recruiting participants. We would like to thank Phoebe Foster for proofreading this article. Finally, our warmest thanks go to all the participants and their families.

## Consent Statement

All human subjects provided informed consent.

## Footnotes

a Abbreviations: AD = Alzheimer’s disease, aAD = amnestic AD, ADHD = attention deficit hyperactivity disorder, ASD = autism spectrum disorder, bvFTD = behavioral frontotemporal dementia, EFA = exploratory factor analysis, FTD = frontotemporal dementia, Fv = focal cortical variants, lvPPA = logopenic primary progressive aphasia, NDDs = neurodevelopmental disorders, DV = neurodevelopmental vulnerability, nfvPPA = non-fluent/agrammatic primary progressive aphasia, rtvFTD = right temporal variant of frontotemporal dementia. svPPA = semantic primary progressive aphasia, Tv = typical variants.

b The matching procedure was performed with the package MatchIt on R studio, using the Nearest Neighbor Matching method with replacement.

a K-means clustering was performed on R Studio with the packages Factoextra, FactoMineR and cluster.

