## Supplementary material for "Neurodevelopmental vulnerability in Alzheimer’s disease and frontotemporal dementia"

|  | **Alzheimer’s disease (AD)** | **Frontotemporal dementia (FTD)** |
| --- | --- | --- |
| **Typical variants (Tv)** | -Amnestic AD (n=38) | -bvFTD (n=18) |
| **Focal cortical variants (Fv)** | -PPA (n=6)  -Frontal variant (n=5)  -Posterior variant (n=2) | -PPA (n=9)  -rtvFTD (n=1)  -Unclassified FTD* (n=5) |

Supplementary Table 1: Classification of typical and focal cortical variants of Alzheimer’s disease and Frontotemporal dementia.

bvFTD: behavioral variant of FTD, PPA: primary progressive aphasia, rtvFTD: right temporal variant of FTD. *Five participants with FTD had an initial amnestic presentation.

|  |  | Group | |  |  |
| --- | --- | --- | --- | --- | --- |
|  |  | Patients who refused (n=21) | Included patients  (n=84) | Statistic | p-value |
| Level of education (years of schooling) | Median (IQR) | 11.0 (1.0) | 11.5 (3) | W = 948 | 0.591^†^ |
|  | Min-Max | 5-20 | 5-20 |  |  |
| MMSE score | Median (IQR) | 23.0 (5.3) | 23.0 (4.0) | W = 813 | 0.826^†^ |
|  | Min-Max | 11-28 | 18-29 |  |  |
| Sex ratio | Number of males (%) | 13 (62) | 45 (54) | χ²= .195 | 0.659^‡^ |
| AD vs FTD ratio | AD/FTD (%/%) | 8/9*  (47/53) | 51/33  (61/39) | χ²= .596 | 0.440^‡^ |
| Subgroups ratio | Fv/Tv  (%/%) | 6/12*  (33/67) | 28/56  (33/67) | χ²= 0 | 1^‡^ |
| Genetic cases (%) | Number of genetic cases (%) | 0 (0) | 8 (.095) | χ²= 1.023 | 0.312^§^ |
| Age at onset (years) | Median (IQR) | 70.1 (7.1) | 65.0 (12.0) | W = 643 | 0.153^†^ |
|  | Min-Max | 51.6-76.3 | 43-80 |  |  |
| Age at diagnosis (years) | Median (IQR) | 72.5 (6.7) | 71.4 (12.1) | W = 836 | 0.716^†^ |
|  | Min-Max | 58.0-77.2 | 43.8-87.9 |  |  |
| Age (years) | Median (IQR) | 74.1 (8.1) | 72.9 (12.6) | W = 864 | 0.889^†^ |
|  | Min-Max | 60.6-78.1 | 45.0-88.3 |  |  |
| Diagnosis timeframe (years) | Median (IQR) | 2.2 (2.3) | 3 (2.6) | W = 1028 | 0.058^†^ |
|  | Min-Max | 0.1-9 | 0.4-14.3 |  |  |
| Disease duration (years) | Median (IQR) | 1.8 (2.2) | 1.6 (2.3) | W = 879 | 0.984^†^ |
|  | Min-Max | 0.0-5.8 | 0.0-17.8 |  |  |

Supplementary Table 2: Demographic description of patients who refused to take part in the protocol.

Fv: focal variants, Tv: typical variants. *One patient had nfvPPA with undefined physiopathology. Three other patients were incorrectly screened and had Lewy Body Dementia. ^†^Bilateral Mann-Whitney test.

Neurodevelopmental disorders screening questionnaire

**NAME:**

**First name:**

**Date of Birth:**

**Sex:**

- Female
- Male

**Have you been diagnosed with a neurodevelopmental disorder? If so, which one(s)?**

**Education**

1) Have you repeated a year and/or been in an adapted class?

- Yes
- No

If yes, please specify:

2) Did you need help learning to read write, count or draw in primary school?

- Private tutoring with a teacher
- Frequent practice with parents or close adults
- Special needs assistant, Individual Educational Success Plan, Tailored program, etc.?
- Part-time
- Other adaptations

3) Did you receive medical or paramedical support, such as:

- Speech and language therapy
- occupational therapy
- psycho-motor therapy
- psychologist
- child psychiatrist
- neuropsychologist

4) What is your highest level of education or most recent diploma?

5) What do you do for a living?

6) Are you:

- right-handed
- left-handed
- ambidextrous

***For each of the following questions, rate the intensity of the behavior mentioned between 0 (no difficulty) and 5 (extremely marked or frequent difficulty):***

***Part 1: When you were a child:***

1) Did you have school phobia? Did you hate school? Was it extremely difficult or even impossible to attend?

0	1	2	3	4	5

2) Did you reverse letters (e.g., writing 'hosre' instead of 'horse') or numbers (e.g., '58' instead of '85')?

0	1	2	3	4	5

3) Were you awkward or uncomfortable moving in space?

0	1	2	3	4	5

4) Did you have trouble managing your emotions (e.g., anger outbursts) or stress?

0	1	2	3	4	5

5) Did you have issues with eating, such as selective eating or an eating disorder?

0	1	2	3	4	5

6) Did you struggle finding your bearings or keeping track of time?

0	1	2	3	4	5

7) Did you have trouble remembering or learning new things?

0	1	2	3	4	5

8) Did you have difficulties in reading, being slower or making more mistakes than your peers?

0	1	2	3	4	5

9) Did you feel you were different from the others, out of step?

0	1	2	3	4	5

10) Did those around you tell you that you were inattentive, daydreaming, or that you lacked concentration?

0	1	2	3	4	5

11) Have you encountered any difficulties in your relationships with family, friends or school mates (harassment, etc.)?

0	1	2	3	4	5

12) Did you want to be the first to answer questions, not waiting for the end of the questions or interrupting?

0	1	2	3	4	5

13) Did you have difficulties with spelling (dictation)?

0	1	2	3	4	5

14) Did you find it difficult to organize your daily routine (homework, tidying up, etc.)?

0	1	2	3	4	5

15) Did you have trouble taking notes in class?

0	1	2	3	4	5

16) In primary and secondary school, did you have any difficulties with mental arithmetic (i.e., without using your fingers to count)?

0	1	2	3	4	5

17) Did you have difficulties acquiring speech or fluency: learning to speak, stuttering, articulation or pronunciation difficulties?

0	1	2	3	4	5

18) Did you struggle mastering certain gestures like buttoning clothes, tying shoelaces, brushing your teeth, using scissors, holding cutlery?

0	1	2	3	4	5

19) Did you have trouble learning your multiplication tables?

0	1	2	3	4	5

20) Did you have trouble learning to write and form letters?

0	1	2	3	4	5

21) Did you have trouble finding your bearings in space?

0	1	2	3	4	5

22) Did you have comments about the “poor appearance” of your notebooks, homework or assignments?

0	1	2	3	4	5

23) Did you find it hard to sit still? Were you told you were “unruly and restless”?

0	1	2	3	4	5

24) In primary and secondary school, did you have any difficulties with geometry (drawing figures, etc.)?

0	1	2	3	4	5

25) Have you had trouble learning to swim, play with a ball, play with a racket, rollerblade or ride a bike?

0	1	2	3	4	5

26) Did you make any careless mistakes?

0	1	2	3	4	5

27) Did you find it difficult to make friends, fit in with your classmates, or take part in extracurricular activities?

0	1	2	3	4	5

***Part 2: As an adult (before the first symptoms of neurodegenerative disease):***

***For each of the following questions, rate the intensity of the behavior mentioned between 0 (no difficulty) and 5 (extremely marked or frequent difficulty):***

28) Are you experiencing difficulties in your professional relationships?

0	1	2	3	4	5

29) Do you tend not to tie/untie your shoe laces?

0	1	2	3	4	5

30) Are you less comfortable than your peers when it comes to reading? For example, do you find it difficult to read movie subtitles?

0	1	2	3	4	5

31) Do you have trouble completing two tasks at the same time?

0	1	2	3	4	5

32) Do you consider yourself “unstable” in romantic relationships?

0	1	2	3	4	5

33) Do you have trouble understanding complex verbal instructions?

0	1	2	3	4	5

34) Do you have trouble putting things in order when doing something that requires organization?

0	1	2	3	4	5

35) Do you have trouble managing your emotions?

0	1	2	3	4	5

36) Do you consider yourself “unstable” professionally (for example, do you frequently change jobs)?

0	1	2	3	4	5

37) Do you find it difficult to finalize the last details of a project once you have achieved the most rewarding parts?

0	1	2	3	4	5

38) Do you have difficulties understanding others' emotions, humor, or implied meanings?

0	1	2	3	4	5

39) Do you have difficulties with spelling?

0	1	2	3	4	5

40) Do you have trouble remembering appointments or obligations?

0	1	2	3	4	5

41) Do you have trouble taking notes, in meetings for example?

0	1	2	3	4	5

42) Do you struggle to eat neatly if you're not paying attention?

0	1	2	3	4	5

43) Do you ever feel overly active and compelled to do something, as if you were driven against your will by a motor?

0	1	2	3	4	5

44) When something requires a lot of thought, do you ever avoid doing it or put it off?

0	1	2	3	4	5

45) Do you ever fidget or wiggle your hands or feet when you have to sit still for long periods?

0	1	2	3	4	5

46) Do you misplace items (keys, documents, mobile phone, etc.?)

0	1	2	3	4	5

**CHILD SCORE:**

**ADULT SCORE:**

**TOTAL SCORE:**

Supplementary Figure 1: English version of the questionnaire investigating retrospective neurodevelopmental disorders symptoms.

|  | **Measurement statistical analysis (MSA)** |
| --- | --- |
| **General** | 0.692 |
| **score_1** | 0.771 |
| **score_2** | 0.543 |
| **score_3** | 0.659 |
| **score_4** | 0.525 |
| **score_6** | 0.648 |
| **score_7** | 0.845 |
| **score_8** | 0.736 |
| **score_9** | 0.584 |
| **score_10** | 0.715 |
| **score_11** | 0.679 |
| **score_12** | 0.578 |
| **score_13** | 0.746 |
| **score_14** | 0.516 |
| **score_15** | 0.823 |
| **score_16** | 0.754 |
| **score_17** | 0.641 |
| **score_19** | 0.571 |
| **score_20** | 0.775 |
| **score_22** | 0.735 |
| **score_23** | 0.534 |
| **score_24** | 0.665 |
| **score_26** | 0.666 |
| **score_27** | 0.606 |

 Supplementary Table 3: Kaiser-Meyer-Olkin test for Sampling Adequacy.

Scores refer to items of the childhood part of the questionnaire.

|  | Factor | | | | | |  | Factor interpretation | Cumulative variance (%) |
| --- | --- | --- | --- | --- | --- | --- | --- | --- | --- |
| Item | 1 | 2 | 3 | 4 | 5 | 6 | Uniqueness |  |  |
| 1 | 0.771 |  |  |  |  |  | 0.373 | General academic struggles | 13.9 |
| 7 | 0.764 |  |  |  |  |  | 0.314 |  |  |
| 15 | 0.599 |  |  |  |  |  | 0.427 |  |  |
| 26 | 0.533 |  |  |  |  |  | 0.503 |  |  |
| 20 | 0.432 |  |  |  |  |  | 0.725 |  |  |
| 6 | 0.415 |  |  |  |  |  | 0.750 |  |  |
| 8 |  | 0.775 |  |  |  |  | 0.257 | Reading | 20.7 |
| 27 |  |  | 0.784 |  |  |  | 0.389 | Psycho-social abilities | 27.5 |
| 9 |  |  | 0.639 |  |  |  | 0.530 |  |  |
| 11 |  |  | 0.523 |  |  |  | 0.415 |  |  |
| 14 |  |  |  | 0.785 |  |  | 0.390 | Executive planning | 33.9 |
| 23 |  |  |  |  | 0.711 |  | 0.487 | Behavioral inhibition | 40.2 |
| 12 |  |  |  |  | 0.610 |  | 0.628 |  |  |
| 17 |  |  |  |  |  | 0.637 | 0.346 | Specific learning skills | 44.5 |
| 16 |  |  |  |  |  | -0.427 | 0.600 |  |  |

Supplementary Table 4: Exploratory Factor Analysis of the childhood items of the questionnaire.

Extraction method: Principal axis; Rotation method: Oblimin. Items designate the number of the question found in the questionnaire. Items 10, 13, 3, 2, 22, 19, 4, 24 did not contribute to factors (< to the .4 cut-off).

|  | **1** | **2** | **3** | **4** | **5** | **6** |
| --- | --- | --- | --- | --- | --- | --- |
| **1** | — | 0.365 | 0.197 | 0.3087 | 0.1714 | 0.1407 |
| **2** |  | — | 0.161 | 0.1871 | 0.0438 | 0.1274 |
| **3** |  |  | — | 0.0977 | 0.1812 | -0.0653 |
| **4** |  |  |  | — | 0.1388 | -0.0417 |
| **5** |  |  |  |  | — | -0.0161 |
| **6** |  |  |  |  |  | — |

 Supplementary Table 5: Inter-factor correlations of the Exploratory Factorial Analysis.

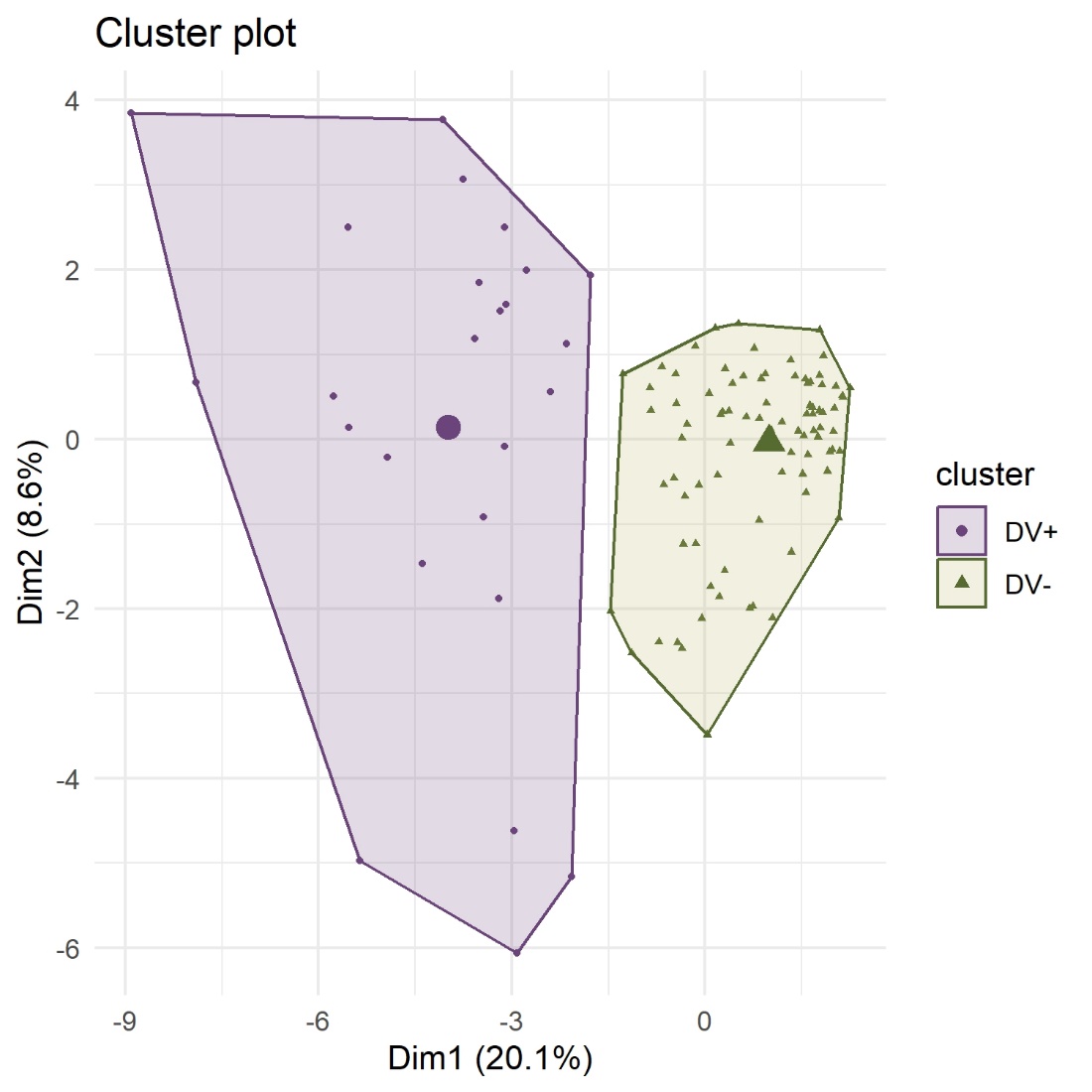

Supplementary Figure 2: Clusters visualization on the first two dimensions.

Enlarged symbols indicate clusters centroids.

|  | **Centroids** | |  |
| --- | --- | --- | --- |
| **Factors** | **Cluster 1** | **Cluster 2** | **p-value** |
| General academic struggles | 1.294 | -0.261 | **<.001** |
| Reading | 1.174 | -0.237 | **<.001** |
| Psycho-social abilities | 1.014 | -0.205 | **<.001** |
| Executive planning | 1.131 | -0.228 | **<.001** |
| Behavioral inhibition | 0.380 | -0.077 | 0.040 |
| Specific learning skills | 0.427 | -0.086 | 0.090 |

Note: Hₐ μ 1 > μ 2

Supplementary Table 6: Centroids of clusters across factors.

Unilateral Mann-Whitney test showed statistically higher mean values in Cluster 1 for all factors except Behavioral inhibition and Specific learning skills.
